# GLP-1-receptor agonist use is associated with shorter survival in patients with amyotrophic lateral sclerosis and diabetes mellitus: a multicenter retrospective study

**DOI:** 10.1101/2025.05.05.25326993

**Authors:** Ikjae Lee, Jiyoon Hwang, Erica Scirocco, Sabrina Paganoni, Nan Jiang, Keith Pearson, Stephanie Dobak, Hristelina Ilieva, Yuyao Sun, Edward Kasarskis, James Russell, Sonya Dunlap, Michael Baer, Colin Quinn, Andrea Charvet, Eduardo Locatellli, Ramita Karra, Martina Wiedau, Yigit Karasozen, Gordon Jewett, Angela Russel, Jonathan Tiu, Robert Connors, Sarah Breevoort, Katherine Ruzhansky, Mehdi Ghasemi, Sorieba Fofanah, Mark Garret, Elijah Stommel, Seonjoo Lee, Kathleen Stolwyk, Matthew Harms, Neil A. Shneider, Michael Rosenbaum

**Author notes:** Corresponding author: Ikjae Lee, MD MSc, Address: 710 W 168th street, NI-3, New York, NY 10032. Statistical analysis performed by: Ikjae Lee. Search terms: 1. Amyotrophic Lateral Sclerosis 2. GLP-1 3. Survival 4. Progression 5. Diabetes Mellitus.

## Abstract

**Background:** Glucagon-like peptide-1receptor agonists (GLP-1RA) are increasingly used to treat diabetes mellitus (DM) and obesity. However, their impact on the progression of amyotrophic lateral sclerosis (ALS) among those with comorbid DM is unknown.

**Objective:** We examined the relationship between GLP-1RA treatment and disease progression among people with ALS and DM.

**Methods:** This multicenter retrospective chart review study used an electronic health record search at 14 institutions to identify consecutive patients with a confirmed ALS diagnosis and documented comorbid type 1 or type 2 DM. All charts were reviewed for demographics, disease history, medication use, and tracheostomy/survival status. Patients who did not meet Awaji ALS diagnostic criteria, lacked a documented history of DM, or had insufficient records were excluded. Tracheostomy-free survival was compared between the GLP-1RA group (exposed) and No-GLP-1RA group (unexposed) using Kaplan-Meier survival curves and Cox proportional hazards models adjusted for age and region of onset (bulbar, respiratory, and diffuse, versus spinal).

**Results:** A total of 308 ALS patients with comorbid DM were included from 14 institutions, of whom 39 (13%) were treated with one or more GLP-1RA. In the GLP-1RA group, diagnostic delay was shorter (10.2 vs 15.9 months, p<0.001), and revised ALS Functional Rating Scale (ALSFRS-R) score decline from symptom onset to first measure was faster (1.5 vs 1.1 points/month, p=0.023) compared with the No-GLP-1RA group. Other baseline characteristics, including age, sex, race, region of onset, and first ALSFRS-R score, were not significantly different between groups. Tracheostomy-free survival from symptom onset was numerically shorter in the GLP-1RA group (median survival 27.0 vs 36.7 months; log-rank p=0.073). After adjusting for age and region of onset, GLP-1RA exposure was associated with an increased risk of tracheostomy or death (hazard ratio 1.56, 95% confidence interval 1.06-2.28, p=0.024).

**Conclusions:** In this multicenter cohort, GLP-1RA treatment was associated with faster disease progression and shorter tracheostomy-free survival in people with ALS and comorbid DM.

## Introduction

Amyotrophic lateral sclerosis (ALS) is a fatal neurodegenerative disease characterized by progressive muscle weakness, leading to death in two to five years after symptom onset.^1^ In the United States, approximately 5,000 new cases of ALS are diagnosed annually.^2^ Despite the availability of various ALS treatments and ongoing research into new therapies, no cure currently exists to stop or reverse the progression of this disease.^3^

Low adiposity and weight loss are common in ALS, and they are known risk factors for faster disease progression and shorter survival.^4-8^ Current ALS treatment guidelines recommend stabilization of weight through gastrostomy and high-caloric supplements.^9^ Recent observations also indicate that diets with higher glycemic index and load are associated with slower functional decline and longer survival,^10,11^ suggesting the importance of adequate dietary glycemic responses. In this context, the recent rise of glucagon-like peptide-1 receptor agonist (GLP-1RA) use reaching over 15% of United States (US) adult population is concerning.^12^

GLP-1RAs are potent synthetic analogs of endogenous GLP-1, with long-lasting action through higher binding affinity for albumin and resistant to degradation by dipeptidyl peptidase 4 (DPP4) enzyme.^13^ Through acting on receptors distributed throughout peripheral organs and the brain, GLP-1RA slows gastric emptying, reduces appetite and hepatic glucose production, resulting in lower food intake, slower nutrient absorption, lower blood glucose levels, and weight loss.^13^ Although weight loss and lower adiposity are associated with cardiovascular benefits in the general population,^14^ it has been linked to rapid disease progression among the ALS population.^4,5,8^ With the increasing use of GLP-1RAs, it is likely that a growing number of ALS patients are being treated with these medications, raising concerns about their potential impact on ALS outcomes. We reported recently that a person with ALS and type 2 DM (T2DM) progressed rapidly after the initiation of semaglutide, a commonly prescribed GLP-1RA.^15^ However, whether semaglutide treatment affected the course of disease or the temporal relationship was coincidental is unknown.

In this multicenter retrospective chart review study spanning 14 institutions, we examined frequency and patterns of antihyperglycemic medication prescription and investigated differences in survival rates between those with and without GLP-1RA treatments.

## Methods

This was a multicenter retrospective chart review study conducted across 14 institutions. Institutional review board (IRB) approval was obtained at each participating institution (lead site IRB# AAAV2579). Electronic health records were screened for ALS (ICD-10 G12.21) and DM (ICD-10 E08–E13) diagnostic codes or chart documentation. Patients were included if they were over 18 years old, had a confirmed ALS diagnosis, met the Awaji ALS diagnostic criteria,^16^ diagnosed ALS between Jan/2020 and Dec/2023, and had documented comorbid type 1 or type 2 DM. Those who did not meet these criteria or had undergone tracheostomy before their first clinic encounter were excluded. For patients who met inclusion and exclusion criteria, detailed chart reviews were conducted to extract data on demographics, anthropometric measures, disease history, functional assessments, medication use, tracheostomy status, and survival.

Demographic variables included reported sex (male vs. female), race (White vs. Asian vs. Black vs. Other vs. Declined), and age at symptom onset. Anthropometric measures included height and weight at diagnosis and follow up. Body mass index (BMI) was calculated from height and weight. Disease history included the region of onset (spinal vs bulbar vs respiratory vs diffuse), diagnostic delay, first documented revised ALS functional rating scale (ALSFRS-R) total score, ALS medications (riluzole, edaravone, tofersen), and presence of pathogenic variants. Diagnostic delay was calculated as the difference, in days, between the date of ALS diagnosis and the date of symptom onset. If symptom onset was reported in month and year, 15 was imputed for the day. The baseline rate of progression was calculated using the following formula: (48 – first measured ALSFRS-R)/disease duration from symptom onset to first ALSFRS-R measurement (months). Medications were reviewed, focusing on the DM medications. The name, first documented date, and last documented date of the DM medications were obtained. Survival and tracheostomy status with dates were recorded. If death was not confirmed, the last known survival date was recorded.

### List of DM Medications

Insulin: Insulin glulisine (Apidra®), Insulin aspart (Novolog®), Insulin lispro U-100/U-200 (Humalog®), Regular insulin (Novolin R, Humulin R), NPH insulin (Novolin N, Humulin N), Insulin detemir (Levemir®), Insulin U-100 (Lantus®, Basaglar®), Insulin glargine U-300 (Toujeo®), Insulin degludec U-100/U-200 (Tresiba®)

Metformin (or containing metformin): ActoPlus Met®, ActoPlus Met XR®, Avandamet®, Fortamet®, Glucophage®, Glucophage XR®, Glucovance®, Glumetza®, Invokamet®, Janumet®, Janumet XR®, Jentadueto®, Kazano®, Kombiglyze XR®, Metaglip®, PrandiMet®, Riomet®, Xigduo XR

GLP1 agonists: Dulaglutide (Trulicity®). Exenatide (Byetta®). Exenatide extended-release (Bydureon®). Liraglutide (Victoza®). Lixisenatide (Adlyxin®). Semaglutide injection (Ozempic®, Weagovy®). Semaglutide tablets (Rybelsus®), Tirzepatide (Mounjaro®, Zepbound®).

DPP4 inhibitors: sitagliptin (Januvia®, Zituvio®), sitagliptin+metformin (Janumet®), linagliptin (Tradjenta®), alogliptin (Nesina®, Vlpidia®), Saxagliptin (Onglyza®)

Sodium-glucose cotransporter 2 (SGLT2) inhibitors: dapagliflozin(Farxiga®, Forxiga®), empagliflozin (Jardiance®), canagliflozin(Invokana®), ertugliflozin(Steglatro®)

PPAR-γ agonists: pioglitazone (Actos®), rosiglitazone (Avandia®)

Sulfonylurea: glyburide(Glynase®), glimepiride(Amaryl®), glipizide (Glucotrol®)

Meglitinide: repaglinide (Prandin®)

### Statistical Analysis

Baseline characteristics were summarized and compared between ALS patients who received GLP-1RA and those without such exposures using T-tests for continuous variables and Chi-square tests for categorical variables. The Kaplan-Meier survival curves and log-rank tests were used to compare tracheostomy-free survival from symptom onset between participants with and without exposure to GLP-1RA. Cox proportional hazard models were used to compare survival between groups, adjusted for covariates. Covariates included age and region of onset (bulbar, respiratory, and diffuse, versus spinal). In sensitivity analysis, survivals were compared between groups based on other classes of DM medications: DPP4 inhibitors; biguanides; insulin; sulfonylurea; sodium-glucose cotransporter 2 (SGLT2) inhibitors; PPAR-γ agonist; meglitinide.

Statistical analyses were conducted using R version 4.4.1., a p-value less than 0.05 was considered significant.

## Results

A total of 308 unique ALS patients with comorbid DM were included across 14 institutions, with a mean age of 65.9 years and 57% male. The most commonly prescribed class of DM medication was biguanides (65%), followed by insulin (36%), sulfonylurea (19%), SGLT2 inhibitors (16%), DPP4 inhibitors (16%), GLP-1 receptor agonists (13%), PPAR-γ agonists (5%), and meglitinide (1%).

Among 308 patients, 39 (13%) were treated with one or more GLP-1RA, while 269 (87%) were never exposed (No-GLP-1RA group). The mean age was 64.8 years in the GLP-1RA group and 66.0 years in the No-GLP-1RA group, with the difference not reaching statistical significance (p=0.44). The GLP-1RA group had a shorter diagnostic delay compared to the No-GLP-1RA group (10.2 vs 15.9 months, p<0.001) and a faster baseline rate of disease progression (1.5 vs 1.1 points per month, p=0.025). Age, sex, race, region of onset, BMI at diagnosis, first documented ALSFRS-R total score, and frequencies of riluzole and edaravone treatment were not significantly different between groups. [Table 1]

**Table 1.** Baseline characteristics of ALS patients with and without the use of GLP-1RA (N=308)

|  | GLP-1RA (n=39) | No-GLP-1RA (n=269) | P-value |
| --- | --- | --- | --- |
| Age at symptom onset, mean (SD) | 64.8 (9.2) | 66.0 (9.5) | 0.44 |
| Sex, M (%) | 23 (59%) | 154 (57%) | 0.99 |
| Race, n (%) | White 26 (66.7%), African American 4 (10.3%), Asian 1 (2.6%), Other 8 (20.5%) | White 168 (62.5%), African American 27 (10.0%), Asian 22 (8.2%), Other 52 (19.3%) | 0.88 |
| Type 1 DM (%) | 0 (0%) | 24 (9%) | 0.11 |
| Bulbar onset (%) | 9 (23%) | 80 (30%) | 0.53 |
| Spinal onset (%) | 30 (77%) | 166 (62%) | 0.10 |
| Respiratory onset (%) | 0 (0%) | 12 (4%) | 0.37 |
| Diffuse onset (%) | 0 (0%) | 10 (4%) | 0.46 |
| Diagnostic delay (months) | 10.2 (7.3) | 15.9 (12.8) | <0.001 |
| BMI at diagnosis | 28.0 (5.4) | 27.3 (5.4) | 0.49 |
| First documented ALSFRS-R | 32.0 (8.9) | 33.2 (8.4) | 0.44 |
| Rate of progression | 1.5 (1.0) | 1.1 (1.1) | 0.023 |
| Riluzole (%) | 34 (87%) | 234 (87%) | 1.00 |
| Edaravone (%) | 10 (26%) | 77 (29%) | 0.84 |

GLP-1RA: glucagon-like peptide 1 receptor agonist; SD: standard deviation; M: male; DM: diabetes mellitus; BMI: body mass index; ALSFRS-R: revised amyotrophic lateral sclerosis functional rating scale.

The Kaplan-Meier survival curve for tracheostomy-free survival shows a numerically shorter median survival in the GLP-1RA group compared to the No-GLP-1RA group (27.0 vs 36.7 months; log-rank p=0.073). [Figure 1] In a Cox proportional hazards model adjusted for age and region of onset, the GLP-1RA group was associated with an increased risk of tracheostomy or death (hazard ratio 1.56, 95% confidence interval [CI] [1.06, 2.28], p=0.024).

**Figure 1.**
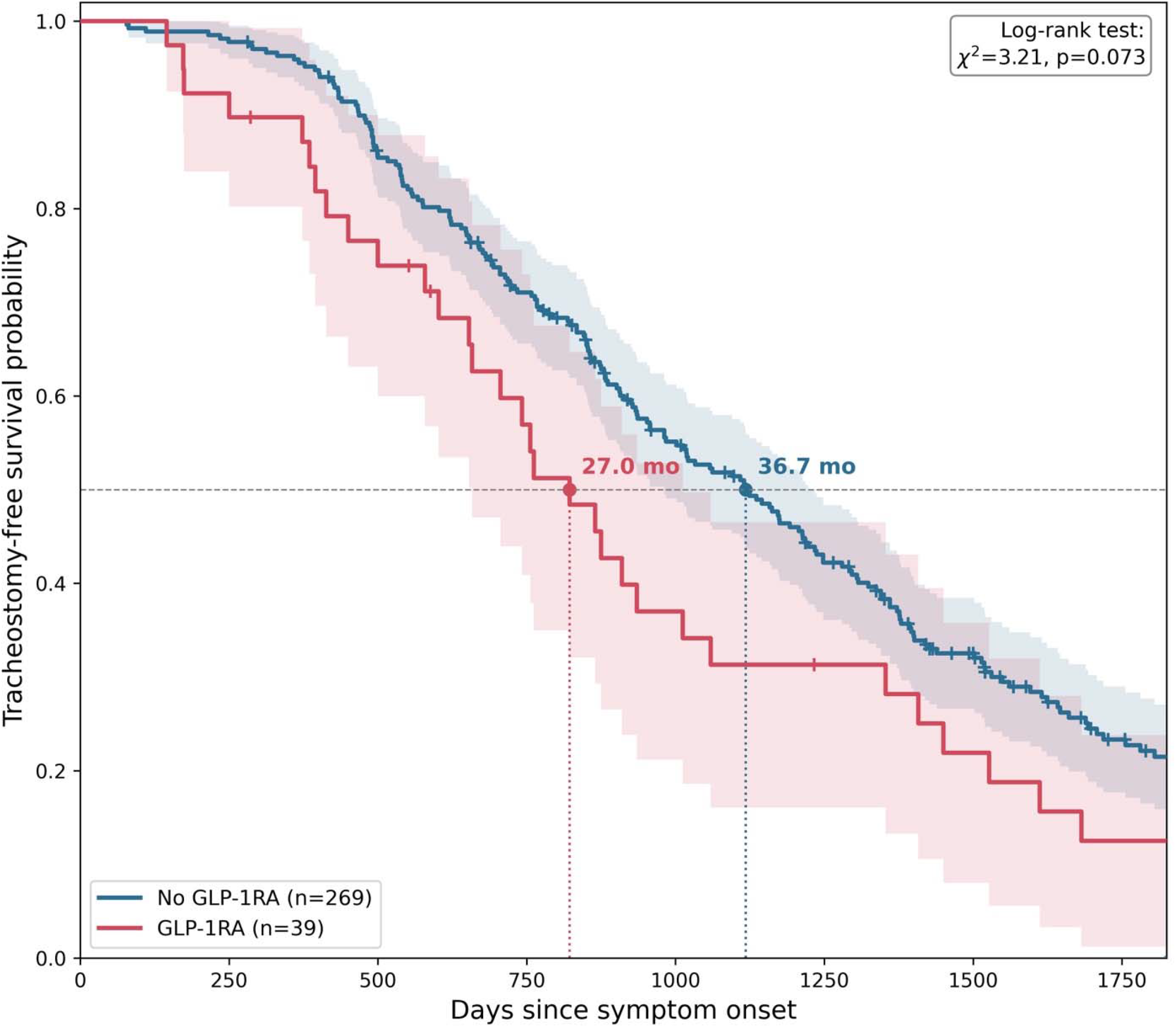
Kaplan-Meier tracheostomy-free survival curve for the multicenter cohort (N=308), comparing GLP-1RA (n=39) vs. No-GLP-1RA (n=269) groups. Adjusted Cox proportional hazards model (age, region of onset): HR=1.56 (95% CI 1.06-2.28), p=0.024.

In sensitivity analyses examining each class of DM medication individually (adjusted for age and region of onset), none of biguanides (HR 0.97, 95% CI [0.74, 1.27]), DPP4 inhibitors (HR 0.92, 95% CI [0.64, 1.33]), SGLT2 inhibitors (HR 1.02, 95% CI [0.73, 1.44]), PPAR-γ agonists (HR 0.99, 95% CI [0.56, 1.74]), sulfonylureas (HR 1.29, 95% CI [0.94, 1.76]), or insulin (HR 1.24, 95% CI [0.95, 1.63]) were significantly associated with tracheostomy-free survival. Meglitinide exposure was associated with shorter survival (HR 3.74, 95% CI [1.35, 10.36], p=0.011), but this estimate is based on only 4 exposed patients and should be interpreted with caution.

## Discussions

In this multicenter retrospective chart review study, we investigated the association between GLP-1RA use and disease progression in people with ALS and DM. We observed that people treated with GLP-1RA had a significantly shorter diagnostic delay and faster rate of disease progression from onset to first-measured ALSFRS-R compared to those who never received such medications, indicators of more aggressive disease progression.^17^ Importantly, those who were exposed to GLP-1RA had 9.7 months shorter survival compared to those who were not exposed. After adjusting for age and region of onset, GLP-1RA exposure was associated with 56% increased risk of tracheostomy or death. Other classes of DM medication individually, including DPP4 inhibitors, none were significantly associated with tracheostomy-free survival, except meglitinide. However, the exposure to meglitinide was very rare (only 4 patients) that the results cannot be interpreted conclusively. This specificity to GLP-1RA, rather than a shared effect across glucose-lowering medications more broadly, suggests that mechanisms more prominent to GLP-1RA—such as its pronounced effects on appetite suppression, weight loss, and gastric emptying—may be more relevant to this association than glycemic control per se. This interpretation is further supported by previous studies reporting that comorbid DM and hemoglobin A1C levels, markers of glycemic control, are not independently associated with survival in ALS.^18,19^

Given that low adiposity and weight loss are established risk factors for faster ALS progression and shorter survival, the association observed in this study might have been mediated through loss of weight and adiposity. Alternatively, ALS patients might be more vulnerable to GLP-1RA induced muscle mass loss and weakness that has been observed among diabetic and obese populations.^20^ However, the current study does not directly measure body composition changes over time and cannot establish causality; prospective studies incorporating longitudinal weight, body composition, and comprehensive neurological examinations and metabolic measurements are needed to test this hypothesis directly.

Our study has limitations. First, as a retrospective analysis, it is subject to potential selection bias and unmeasured confounders. While we adjusted for key covariates, residual confounding cannot be ruled out.

Second, despite the larger multicenter sample compared to prior single-center work, the number of GLP-1RA-exposed patients remained modest, limiting power for subgroup and sensitivity analyses. Finally, while our study highlights an association between GLP-1 use and decreased survival, it does not establish causality. Future studies are urgently needed to confirm or refute our findings and further examine the underlying biological mechanisms driving this relationship, including the impact of GLP-1 therapy on metabolic homeostasis in people with ALS.

Study funding: The authors received no direct funding for this study.

Conflict of interest disclosures: IL received research funding from the National Institute of Health and Spastic Paraplegia Foundation, received consultation fees from Regeneron and Argenx pharmaceuticals. All other authors declare no conflict of interest exists.

## Data Availability

All data produced in the present study are available upon reasonable request to the authors after the publication of the study.

